# Age- and gender-dependent differences in attitudes towards COVID-19 vaccination and underlying psychological processes

**DOI:** 10.1101/2021.05.28.21257954

**Authors:** Toshiko Tanaka, Tsuyoshi Nihonsugi, Fumio Ohtake, Masahiko Haruno

**Author notes:** Correspondance should be addressed to Dr. Masahiko Haruno, Center for Information and Neural Networks, National Institute of Information and Communications Technology, Suita,Osaka 565-0871, Japan.

## Abstract

The most promising way to prevent the explosive spread of COVID-19 infection is to achieve herd immunity through vaccination. It is therefore important to motivate those who are less willing to be vaccinated. To address this issue, we conducted an online survey of 6232 Japanese people to investigate age- and gender-dependent differences in attitudes towards COVID-19 vaccination and the underlying psychological processes. We asked participants to read one of nine different messages about COVID-19 vaccination and rate their willingness to be vaccinated. We also collected their 17 social personality trait scores and demographic information. We found that males 10-20 years old showed the minimum willingness to be vaccinated. We also found that prosocial traits are the driving force for young people, but the motivation in older people also depends on risk aversion and self-interest. Furthermore, an analysis of 9 different messages demonstrated that for young people (particularly males), the message emphasizing the majority’s intention to vaccinate and scientific evidence for the safety of the vaccination had the strongest positive effect on the willingness to be vaccinated, suggesting that the herding effect arising from the “majority + scientific evidence” message nudges young people to show their prosocial nature in action.

## Introduction

Our society has been enormously damaged by the COVID-19 pandemic. For now, the acquisition of herd immunity through vaccination is the most promising way to control the spread of the infection. Indeed, more than half of the population in the UK has already received its first vaccination, and the number of new infections there is decreasing, making it possible to ease restrictions on civic life gradually. Although the supply of vaccines is not sufficient except in a few countries, deep understanding of people’s different attitudes towards COVID-19 vaccination should be useful for the effective implementation of unprecedentedly large vaccination programs in the future.

Previous studies have already investigated whether different individual characteristics, such as age, gender, and living conditions (income, work environment, perception of infection risk, etc.), influence the willingness to be vaccinated (e.g., influenza). Schmid and colleagues [1] reported not only that low risk and low worry about influenza were barriers to vaccine uptake, but also that being white was positively associated with vaccination in the general population. They also reported that older age and living alone were respectively associated with vaccination positively and negatively. Females and lower income were reported to be often negatively associated with vaccination [2,3,4]. Overall, there are mixed reports on gender differences for vaccinations against COVID-19. Notably, most studies arguing that gender differences exist reported females have lower motivation to vaccinate [5]. As for age, younger people are also less motivated to be vaccinated because of their perception of lower risk [6,7]. In the case of COVID-19, about 30% of infected people are asymptomatic, which may contribute to the spread of infection [8,9]. Therefore, it is critically important to promote the vaccination in young people who are less motivated.

To increase their motivation, understanding the psychological processes underlying their low motivation would be helpful, although previous studies mainly focused on direct reasons for refusing vaccination [10]. Several studies reported that personality traits are linked with the acceptance/hesitancy of the vaccination [11,12,13,14,15]. Thus, the identification of psychological factors would contribute to more effective prevention measures; for example, the design of an effective message that appeals to young individuals who are less motivated towards the vaccination.

To address this issue, we conducted an online survey of 6232 Japanese people (15-59 years old) and investigated age- and gender-dependent differences in their attitudes towards COVID-19 vaccination. We asked the participants to read 1 of 9 different (nudge) messages about COVID-19 vaccination (see Methods) and then to report their degree of willingness (Question A) or refusal (Question B) to take the vaccination using a 7-scale forced key choice (1:not at all, 2: hardly at all, 3: not much, 4: neutral, 5: somewhat, 6: much and 7: definitely). We also collected 17 social personality trait scores, including altruistic beliefs, neuroticism, collective responsibility, conscientiousness, and anxiety (see Supplementary Table S1 online), and demographic information from the participants.

We first looked at the effects of demographic factors on the willingness or refusal of the COVID-19 vaccination. We then analyzed willingness and refusal scores by LASSO regression, identified the personality traits contributing to predict those scores at each individual level and investigated how the personality traits important for the prediction are different among different age and gender groups. We finally evaluated the effectiveness of the 9 different nudge messages for different age- and gender-groups to promote willingness and reduce refusal of the vaccination.

## Results

Table 1 displays demographic decompositions of the participants who showed high degrees of willingness or refusal of the COVID-19 vaccination. For willingness (Question A), if a participant responded 1) not at all to 3) not much, we judged the participant had a low likelihood of getting the vaccine (score: -1). On the other hand, if a participant responded 5) somewhat to 7) definitely, we judged the participant had a high likelihood of getting the vaccine (score: +1). Neutral response 4) was scored 0. Similarly, for refusal of the vaccine (Question B), if a participant responded 1) as soon as possible to 3) one year ahead, the participant was regarded as wanting the vaccination immediately (score: -1). On the other hand, response 4) two years to 6) five years ahead were judged as intending to be vaccinated sooner or later (score: 0). If a participant responded 7) never, we judged that the participant would always refuse the vaccination (score: 1). These scores were also used in the subsequent multiple linear regression analysis of the willingness and refusal scores by demographic decomposition. Overall, 68.5% (n = 4270) and 12.0 % (n = 745) of the participants showed high willingness and refusal of the vaccination, respectively. Participants who were young, with no underlying diseases, with a low annual income, with a low level of usual preventive attitude, and working in a workplace not related to healthcare had a significantly lower likelihood of getting a COVID-19 vaccine.

**Table 1.**
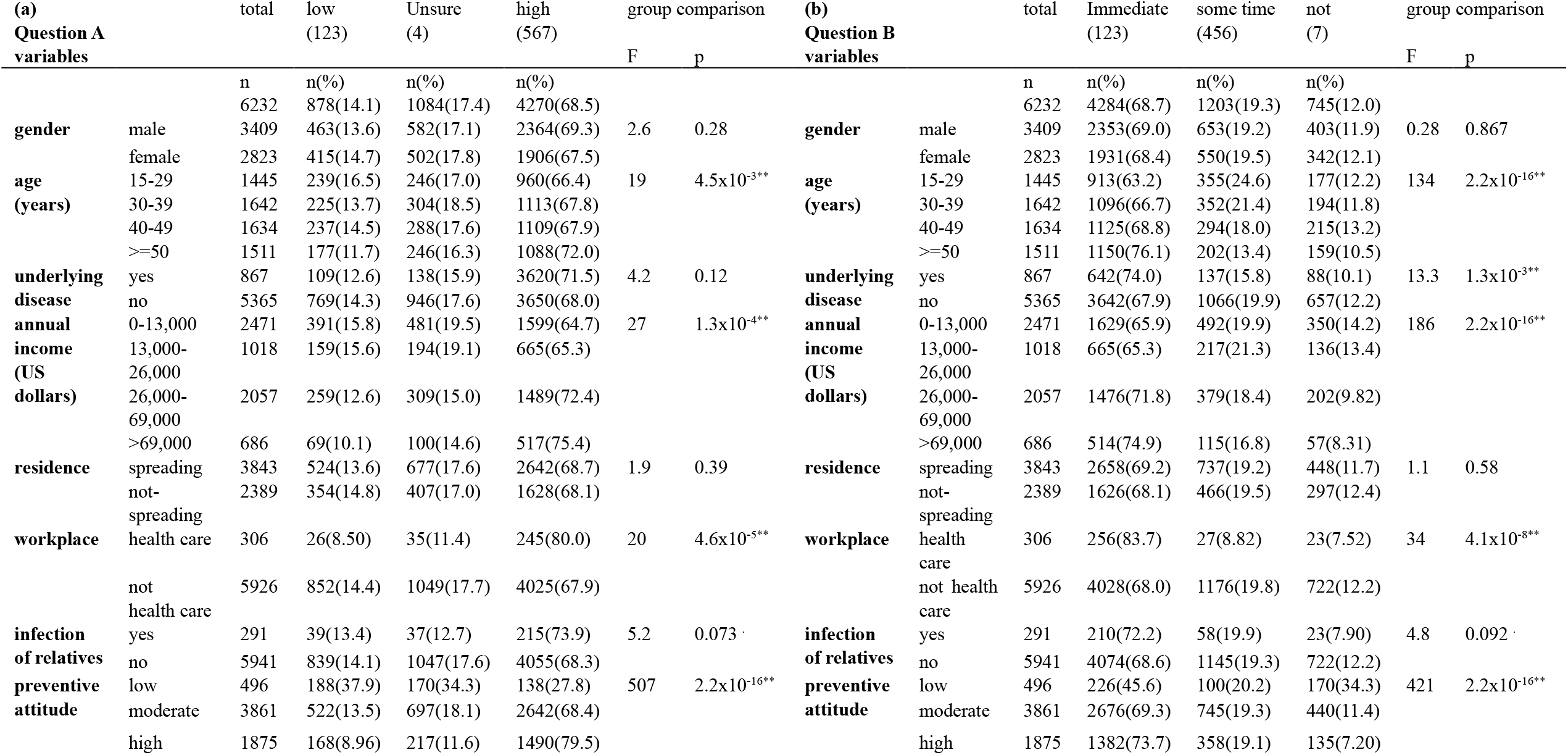
Univariate decomposition of willingness (Question A, Table 1a) and refusal (Question B, Table 1b) to vaccinate into different participant demographic characteristics. Codes for significance: 0 ‘**’, 0.01 ‘*’, 0.05 ‘.’ 0.1

Table 2 summarizes the results of the multiple linear regression of the willingness and refusal scores by demographic information. We found a significant gender difference (lower in females) in the willingness to be vaccinated, consistent with previous reports on gender differences in the level of COVID-19 vaccine acceptance [2,16]. We also found that a younger age, no underlying disease, low annual income, workplace unrelated to healthcare and low-level usual preventive attitude are associated with a significantly lower likelihood of getting a COVID-19 vaccine.

**Table 2.**
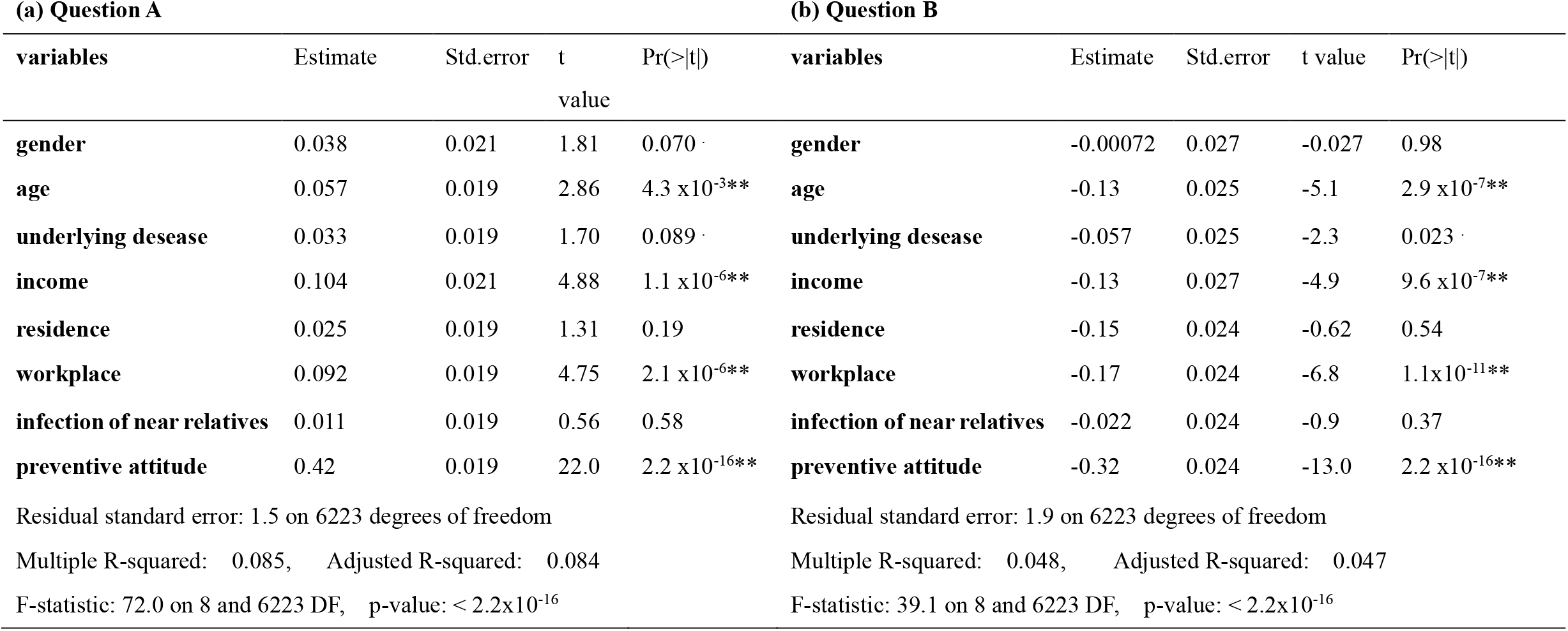
Multiple linear regression of the willingness and refusal to be vaccinated with participants’ demographic characteristics. Codes for significance: 0 ‘**’, 0.01 ‘*’, 0.05 ‘.’ 0.1

Figure 1 visualizes the relative differences in the mean willingness and refusal scores (group mean minus total mean) for age and gender separately. The two-tailed p-values of t-tests between the two groups were calculated and are shown in the lower panels. For the participants’ willingness (Fig. 1a), males in their 50s showed significantly higher motivation towards being vaccinated than males in their 10-20s or 40s. Females in their 40s showed significantly lower motivation than females in their 50s. As for the refusal to be vaccinated (Fig. 1b), males in their 50s showed a significantly lower likelihood of refusal than males in any other age group. Females in their 50s also showed a lower likelihood of refusal than females in their 10-20s. All these results clarified that people (males in particular) in their 10-20s are the least willing and most likely to refuse the vaccination, making a sharp contrast with people in their 50s, who are most willing and least likely to refuse the vaccination.

**Figure 1.**
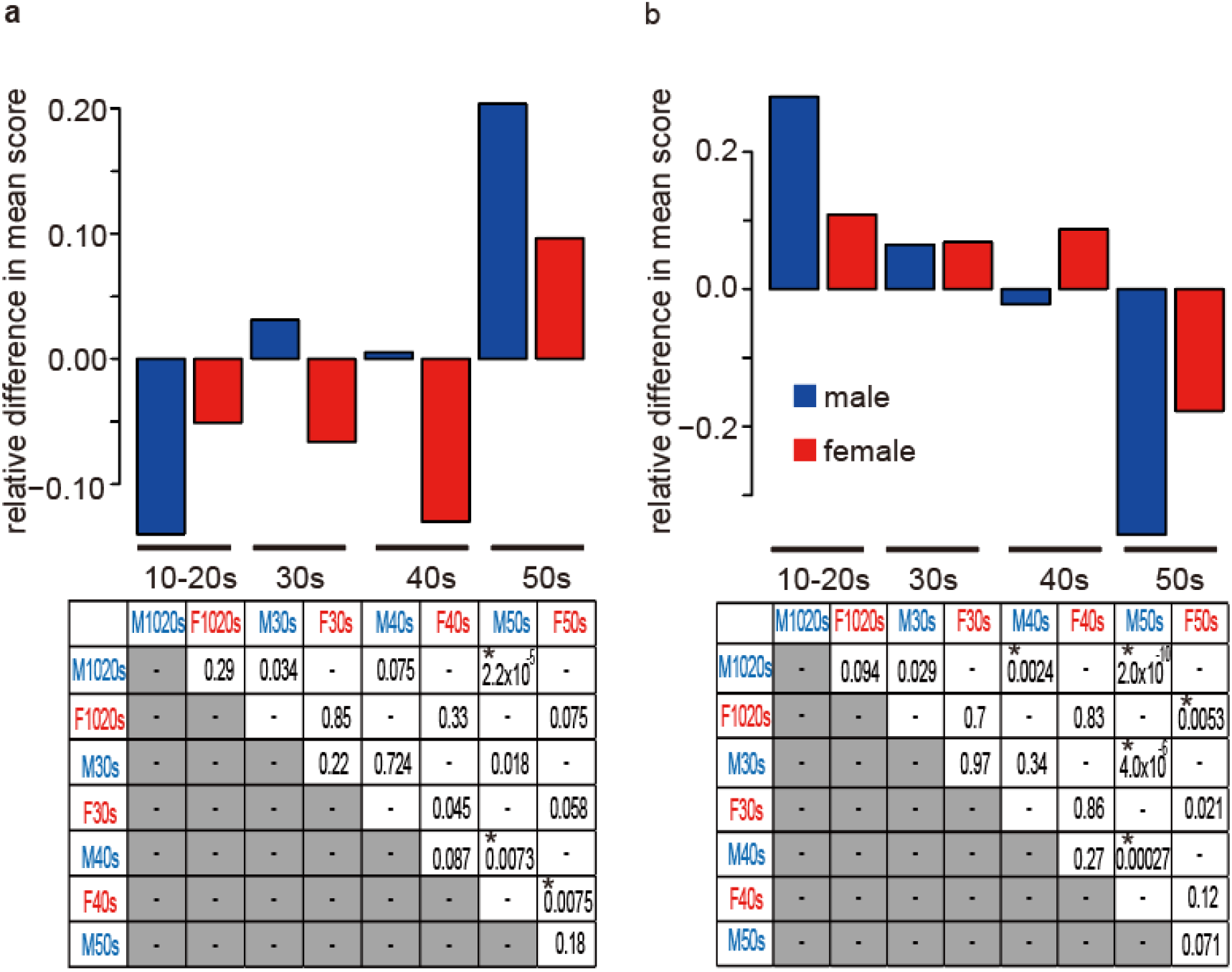
Age- and gender-dependent differences in willingness (a) and refusal (b) to be vaccinated. The upper panels show relative differences in the mean willingness/refusal (group mean minus total mean) (male; blue, female; red). The lower panels represent p-values of t-tests between the two groups. Significance after the correction for multiple comparisons is indicated by *.

We next explored the underlying psychological processes behind the age- and gender-dependent differences in the willingness and refusal to be vaccinated. To investigate the relationships between social personality trait scores and the willingness to be vaccinated, we conducted a LASSO regression of the 7-scale original responses for Question A. LASSO can deal with a large dimensionality of explanatory variables arising from the interaction terms of variables and select the effective explanatory variables sparsely (weights for many non-effective explanatory variables are estimated to be 0) with non-zero weights (beta values) meaning a significant contribution. As the explanatory variables, all social personality scores and demographic information together with the interactions of social personality trait scores with the age and gender variable were used (see Methods for details).

Table 3 summarize the LASSO results for Question A and Question B. For the willingness, agreeableness (Big5_A) had heavy weights in almost all ages and genders, while altruism (TRU_GSS) had a large weight only for females in their 40s whose Big5_A weight was 0. These results demonstrated that the tendency towards prosocial and empathetic considerations increased the motivation to be vaccinated overall. Importantly, among males, the weight of Big5_A was markedly greater for 10-20s. The weight of prosocial score (SVO_P) was also only large for males in their 10-20s. In females, the weight of trait fairness (TRU_WVS) was greater for 10-20s. All these data clarified that for 10-20s, prosocial consideration rather than their own interest is the major driving force of the willingness to receive the COVID-19 vaccination, and this tendency is stronger in males.

**Table 3.** LASSO regression of the willingness and refusal to be vaccinated by social personality trait scores and demographic characteristics (and their interactions). UD, underlying disease; INF, infected close relative; WP, workplace; area, place of residence; income, annual income.

| (a) | 10-20s |  | 30s |  | 40s |  | 50s |  | (b) | 10-20s |  | 30s |  | 40s |  | 50s |  |
| --- | --- | --- | --- | --- | --- | --- | --- | --- | --- | --- | --- | --- | --- | --- | --- | --- | --- |
|  | male | female | male | female | male | female | male | female |  | male | female | male | female | male | female | male | female |
| LAR | 0 | 0.033 | 0 | 0 | 0 | -0.010 | -0.017 | 0 | LAR | 0 | -0.0087 | 0 | 0 | 0 | 0.016 | 0.0088 | 0 |
| RA | -0.052 | 0.035 | 0 | 0 | 0 | -0.015 | -0.010 | -0.072 | RA | 0 | 0 | 0.044 | 0 | 0.0094 | 0 | 0 | 0.022 |
| TIM | 0 | 0.017 | 0 | 0 | 0 | 0 | 0 | 0 | TIM | 0 | 0 | 0.011 | 0 | 0 | 0 | 0 | 0.027 |
| CCP1 | 0.010 | 0 | 0.041 | 0.048 | 0 | 0 | 0 | -0.042 | CCP1 | 0 | 0 | -0.0049 | -0.0079 | 0 | 0 | 0 | 0.012 |
| GSSWVS | 0 | 0 | 0.0043 | 0 | 0 | 0.070 | 0.087 | 0 | GSSWVS | 0 | 0 | 0 | -0.024 | -0.013 | 0 | -0.016 | 0 |
| WVS | 0 | 0.12 | 0 | 0 | 0.056 | 0.050 | 0 | 0.068 | WVS | 0 | -0.044 | 0 | 0 | -0.0075 | -0.022 | -0.0017 | -0.014 |
| GSS | 0 | 0 | 0.027 | 0 | 0 | 0.11 | 0 | 0 | GSS | 0 | -0.0063 | -0.024 | 0 | -0.00022 | -0.046 | 0 | -0.0025 |
| guilt | 0 | 0 | 0 | 0 | 0 | 0 | 0.019 | 0.046 | guilt | 0 | 0 | 0 | 0 | 0 | 0 | -0.028 | 0 |
| inequity | 0.078 | 0 | 0.039 | 0 | 0 | 0 | 0 | 0 | inequity | -0.0026 | 0 | 0 | 0 | 0 | 0 | 0 | 0 |
| ES_E | 0 | 0 | 0 | 0 | 0 | -0.042 | 0 | 0 | ES_E | 0 | 0 | 0 | 0 | -0.010 | 0 | -0.012 | 0 |
| ES_S | 0 | 0 | 0 | -0.0073 | 0 | 0 | 0 | 0 | ES_S | -0.014 | 0 | 0 | 0.0016 | 0 | 0.0059 | 0 | 0 |
| Big5_E | -0.020 | 0 | 0 | 0 | 0 | 0 | 0 | 0 | Big5_E | 0 | 0 | 0.0089 | -1.4x10<sup>-4</sup> | 0 | 0 | 0.00029 | 0 |
| Big5_A | 0.22 | 0.006 | 0.14 | 0.12 | 0.049 | 0 | 0.078 | 0.063 | Big5_A | 0 | 0 | -0.067 | -0.0023 | -0.033 | 0 | 0 | 0 |
| Big5_C | 0 | 0 | 0 | 0 | 0 | 0 | 0 | 0 | Big5_C | 0 | 0.0017 | 0.0046 | 0 | 0 | 0 | 0 | 0 |
| Big5_N | -0.023 | -0.0038 | 0 | -0.017 | 0 | 0 | 0 | 0 | Big5_N | 0.026 | 0 | 0.0069 | 0.028 | 0 | 0 | 0.014 | 0 |
| Big5_O | 0 | 0 | 0 | -0.0047 | 0 | -0.0066 | 0 | -0.028 | Big5_O | 0.0033 | 0.028 | 0 | 0 | 0 | 0 | 0 | 0.0056 |
| SVO_P | 0.062 | 0 | 0 | 0 | 0 | 0 | 0 | 0 | SVO_P | -0.027 | 7.8x10<sup>-5</sup> | 0 | -0.012 | 0 | 0 | 0 | 0 |
| SVO_I | 0 | 0 | 0 | 0 | 0.010 | 0 | 0 | 0 | SVO_I | 0.00061 | 0 | 0 | 0 | -0.0056 | 0 | 0.0089 | 0 |
| IRI_F | 0 | 0 | 0 | 0 | 0.13 | 0 | 0.073 | 0 | IRI_F | 0 | 0 | 0 | 0 | -0.013 | 0 | -0.024 | 0 |
| IRI_PT | 0 | 0 | 0 | 0 | 0 | 0 | 0 | 0 | IRI_PT | 0 | 0 | 0 | 0 | 0 | 0 | 0 | 0 |
| IRI_EC | 0 | 0 | 0 | 0.027 | 0 | 0.075 | 0 | 0.13 | IRI_EC | 0 | 0 | 0 | -0.0088 | 0 | -0.022 | 0 | -0.0032 |
| IRI_PD | 0 | 0 | 0 | 0 | 0.017 | 0 | 0 | 0.057 | IRI_PD | 0 | 0 | 0 | 0 | -0.015 | 0 | -0.014 | 0 |
| SHS | 0 | 0 | 0 | 0 | 0 | 0 | 0 | 0 | SHS | 0 | 0 | 0.020 | 0 | 0.0060 | 0 | 0 | 0 |
| IQ | 0 | 0.015 | 0.056 | 0.15 | 0.0028 | 0.065 | 0 | 0.035 | IQ | 0 | 0 | -0.017 | -0.045 | 0 | 0 | 0 | -0.027 |
| MVS | 0.013 | 0.0049 | 0 | 0 | 0 | 0 | -0.021 | 0 | MVS | 0 | 0 | 0 | 0 | 0 | 0 | 0 | 0 |
| PSS | 0 | 0 | 0.13 | 0.0030 | 0 | 0.017 | 0 | 0 | PSS | 0 | 0 | -0.065 | 0 | -0.0096 | 0 | 0 | 0 |
| RSS | 0 | 0 | 0 | 0 | 0 | 0 | 0 | 0 | RSS | 0 | 0 | 0 | 0 | 0 | 0 | 0 | 0 |
| STAI_S | 0 | 0 | -0.086 | 0 | 0 | 0 | 0 | 0.0039 | STAI_S | 0 | 0 | 0 | 0.0043 | 0 | 0 | 0 | -0.041 |
| STAI_T | 0 | 0 | 0 | 0 | 0 | 0.0097 | 0 | 0 | STAI_T | 0 | 0 | 0 | 0 | -0.011 | -0.026 | 0 | 0 |
| SES | 0 | 0 | 0 | 0 | 0.023 | 0 | 0.072 | 0 | SES | 0 | 0 | 0 | -0.011 | 0 | 0 | 0 | 0 |
| UD | 0.032 |  |  |  |  |  |  |  | UD | -0.0053 |  |  |  |  |  |  |  |
| INF | 0 |  |  |  |  |  |  |  | INF | -0.0043 |  |  |  |  |  |  |  |
| WP | 0.073 |  |  |  |  |  |  |  | WP | -0.0035 |  |  |  |  |  |  |  |
| area | 0 |  |  |  |  |  |  |  | area | 0 |  |  |  |  |  |  |  |
| income | 0.14 |  |  |  |  |  |  |  | income | -0.055 |  |  |  |  |  |  |  |

The weight of stress score (PSS) was large for males in their 30s, but had a small positive beta-value for females. In addition, the weight of conditional cooperation (CCP) was high for both genders in their 30s. For participants in their 40s, the personality traits with a larger weight included Big5_A, imagining empathy (IRI_F) and fairness (TRU_WVS) for males, and altruism (TRU_GSS), empathetic concern (IRI_EC) and trust (TRU_GSSWVS) for females. These stress-related and other-regarding scores promoted the willingness to be vaccinated in participants in their 30s and 40s, suggesting that the infection and serious illness were not considered to be someone else’s problem. Importantly, this view was also consistent with the positive weight of individualistic traits (SVO_I) for males in their 40s, indicating they regarded vaccination as a measure to protect their own health. In addition, the weight of anxiety (STAI_T) was positive for females in their 40s and (STAI_S) was positive for females in their 50s, revealing a connection with a fear of infection. It is also notable that guilt aversion (guilt), i.e., the tendency to avoid a discrepancy between other’s expectation and actual outcome, had a positive effect on males and females in their 50s.

To focus on the refusal of vaccination, we transformed the answer to Question B into a binary variable consisting of either “never” (response 7, score 1) or “others” (response 1-6, score: -1) and used it as the target variable (see Methods for details). Although the personality traits having positive weights for refusal were less frequent because the overall basic trend was a willingness to be vaccinated, personality traits contributing to refusal differed depending on age and gender (Table 3b). Neuroticism (Big5_N) was likely to lead to refusal in many groups, and for people in their 30s or older, risk aversion (RA) and loss aversion (LAR) were also associated with refusal. Importantly, positive weights for individualistic trait (SVO_I) in males in their 10-20s and for openness (Big5_O) in both genders in their 10-20s indicated that individualistic young people were optimistic about their own infection and thereby did not feel vaccination was necessary if they only thought about themselves. Overall, our results demonstrated that younger people vaccinate mainly for others, while in older people, the contributions of self-regarding considerations increase. In particular, males in their 10-20s has a strong tendency to be vaccinated for others and refused for themselves, resulting in their low willingness to be vaccinated (Fig. 1).

An effective way to prevent the spread of COVID-19 is to increase vaccination rates among young people, especially males, considering their minimum willingness to be vaccinated, as identified above. Nudging has been shown to be a useful measure to increase people’s prosocial behaviors while keeping their freedom of choice [17]. In the context of COVID-19, altruistic messages were shown to be effective at preventing infection [18,19]. However, the differences in vaccination-promoting personality traits in different age and gender groups suggest that the effectiveness of nudge messages also differs between these groups. We therefore investigated the effects of nine different nudge messages (see Appendix 1), including altruistic messages as a control. In brief, nudge 1 emphasized the altruism in gain framing, nudge 2 showed scientific evidence in gain framing, nudge 3 showed scientific evidence in loss framing, nudge 4 = nudge 2 + nudge 1, nudge 5 = nudge 3 + nudge 1, nudge 6 = nudge 2 + altruism in loss framing, nudge 7 = nudge 3 + altruism in loss framing, nudge 8 = nudge 2 + majority, and nudge 9 = nudge 3 + majority.

Figure 2 illustrates the proportion of answers to the questions about willingness (Question A) and refusal (Question B) to the vaccination after reading one of the 9 different nudge messages. As with Table 1, answers 6 and 7 (cyan and blue) indicated strong willingness. Compared to the control altruistic nudge (1), nudge 8 showed an increase in the strong willingness ratio among males in their 10-20s (blue asterisk, p = 0.049). The strong willingness ratio of males in their 30s and 40s was sharply decreased for nudge 2 (black crosses, p =0.041 and 0.023, respectively). By contrast, nudges 2-5 showed an increase in the ratio of strong willingness for females in their 30s (blue asterisk, p = 0.046, 0.0074, 0.039, and 0.00070, respectively) and nudge 7 showed an increase in the ratio of strong willingness for females in their 40s (blue asterisk, p = 0.035). No nudge message decreased the strong motivation to vaccinate. For refusal, answer 7 (black) represented the refusal to be vaccinated. The only message that significantly increased the refusal rate was nudge 2 for males in their 10-20s (black asterisk, p = 0.014). The key message from these results is that nudge message 8, which was designed to emphasize the majority’s vaccination intention in addition to scientific evidence of safety, is effective on males in their 10-20s, while nudge messages 3 and 5, which appealed to loss (prospect of increased infections in the case without vaccination), are effective on females in their 30s. These nudges are likely to be applicable to work on the least motivated groups (i.e., males in their 10-20s and females in their 30s and 40s). By contrast, nudge message 2 is likely least effective on males in their 10-20s.

**Figure 2.**
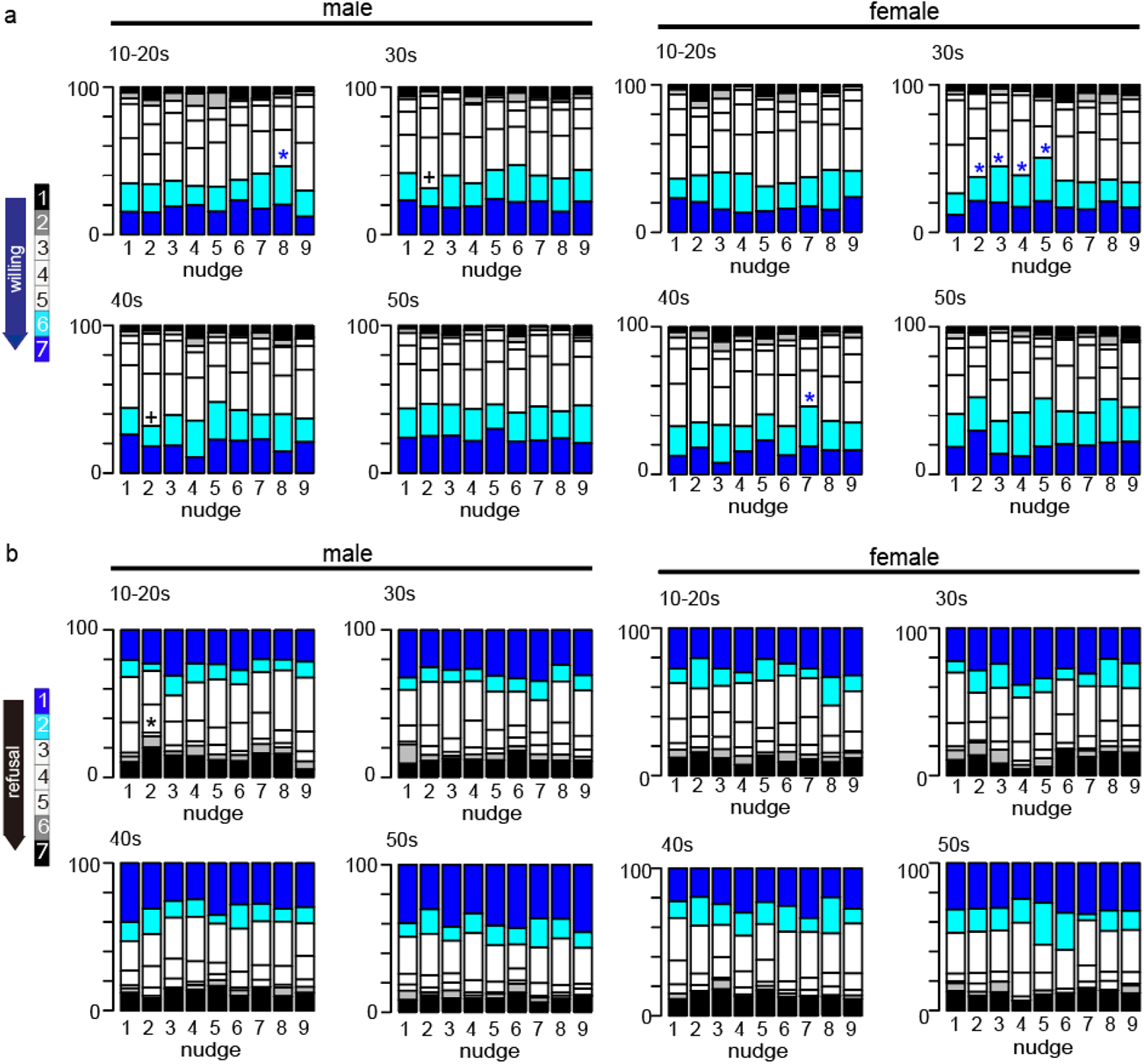
Effect of nudges on willingness/refusal to be vaccinated. The ratios of responses to Question A (a; willingness) and Question B (b; refusal) by age and gender are shown for each nudge. Nudges that significantly increased the willingness to be vaccinated are indicated by blue asterisks, those that decrease the willingness to be vaccinated are indicated by black crosses, and those that promoted refusal to be vaccinated are indicated by the black asterisk.

Next, we assessed the age- and gender-dependent effects of the nudge messages using LASSO regression by incorporating a binary dummy variable representing whether each nudge was sent to a participant or not (see Methods). Figure 3 displays the weights for the interactions between each nudge message and gender and demographic information. For participants in their 10-20s, nudges 8 and 9 promoted willingness in males and females, respectively, while nudge 2 decreased willingness in both genders, consistent with the data in Figure 2. For participants in their 40s, nudges 1 and 5 promoted willingness in males, while nudges 3 and 4 decreased willingness in females and males, respectively (Fig. 3a). Nudges 2 and 6 were shown to promote refusal among males in their 10-20s and 30s, respectively, while nudge 9 reduced refusal among males in their 10-20s. For females, nudge 4 reduced refusal for all ages except 40s.

**Figure 3.**
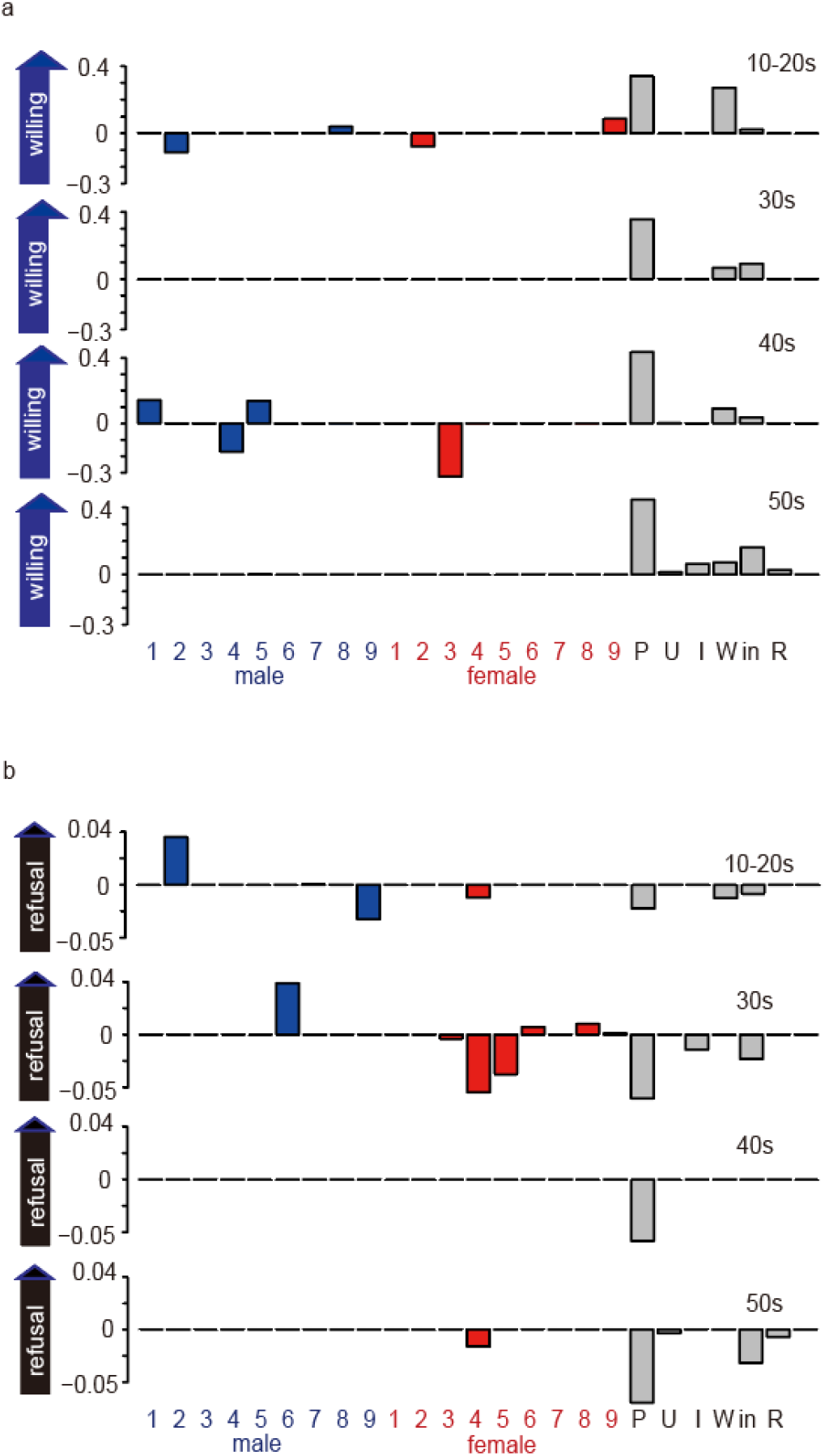
LASSO analysis of the contribution of each nudge on the willingness to be vaccinated for different ages and genders. For each age group, willingness (a) and refusal (b) to be vaccinated were regressed using LASSO, with the following variables as regressors: the interaction between nudge and gender, degree of usual prevention, underlying disease, infection in close relatives, workplace, income, and place of residence.

To summarize, we have so far clarified that 1) nudge 8 was effective at increasing the willingness to be vaccinated among males in their 10-20s, and 2) nudge 2 increased refusal to be vaccinated among males in their 10-20s. It is important to design a nudge-based intervention that does not worsen individual welfare in addition to achieving social goals at a higher rate [20]. Therefore, we confirmed that neither strong discomfort nor resentment was reported from those who received nudge message 8 (see Supplementary Fig. S1 and Table S2 online).

Finally, to directly investigate the psychological processes that account for the effect of nudge 8 on the willingness and refusal to be vaccinated, we conducted a LASSO regression analysis based on the personality and personality-age interaction terms as regressors only for those who received nudge 8 (see Methods section). As shown in Table 4a, the large weights for prosocial orientation (SVO_P) and agreeableness (Big5_A) in the column ‘all (effects of personality traits for all)” suggested that nudge 8 acts on prosocial instinct of all ages. Importantly, the positive weight for agreeableness in people in their 10-20s clarified that this effect is particularly strong in young people. This finding was supported by the positive weight for individualistic trait (SVO_I) for the same group with regards to the refusal to vaccinate (Table 4b).

**Table 4.** LASSO regression of the willingness and refusal to be vaccinated by social personality trait scores and their interactions with age on participants who only received nudge 8. UD, underlying disease; INF, infected close relative; WP, workplace; area, place of residence; income, annual income.

| (a) Question A |  |  |  |  |  | (b) Question B |  |  |  |  |  |
| --- | --- | --- | --- | --- | --- | --- | --- | --- | --- | --- | --- |
| Nudge8 | all | 10-20s | 30s | 40s | 50s | Nudge8 | all | 10-20s | 30s | 40s | 50s |
| LAR | 0 | 0 | 0 | 0 | 0 | LAR | 0 | 0 | 0 | 0 | 0 |
| RA | 0 | 0 | 0 | 0 | 0 | RA | 0 | 0 | 0 | 0 | 0 |
| TIM | 0 | 0 | 0 | 0 | 0 | TIM | 0.0045 | 0 | 0 | 0 | 0 |
| CCP1 | 0 | 0 | 0.022 | 0 | 0 | CCP1 | 0 | 0 | 0 | 0 | 0 |
| GSSWVS | 0 | 0 | 0 | 0 | 0 | GSSWVS | 0 | 0 | 0 | 0 | 0 |
| WVS | 0.018 | 0 | 0 | 0 | -0.089 | WVS | 0 | 0 | 0 | 0 | 0 |
| GSS | 0 | -0.057 | 0 | 0 | 0 | GSS | -0.023 | 0 | 0 | 0 | 0 |
| guilt | 0 | 0.036 | 0 | 0 | -0.078 | guilt | -0.001 | 0 | 0 | 0 | 0 |
| inequity | 0 | 0 | 0 | 0 | 0 | inequity | 0 | 0 | 0 | 0 | 0 |
| ES_E | 0 | 0 | 0 | 0 | 0 | ES_E | 0 | 0 | 0 | 0 | 0 |
| ES_S | 0 | 0 | 0 | 0 | 0 | ES_S | 0 | 0 | 0 | 0 | 0 |
| Big5_E | -0.063 | 0 | -0.11 | 0 | 0 | Big5_E | 0 | 0 | 0 | 0 | 0 |
| Big5_A | 0.11 | 0.21 | 0 | 0 | 0 | Big5_A | 0 | -0.033 | 0 | 0 | 0 |
| Big5_C | 0 | 0 | 0 | 0 | 0 | Big5_C | 0 | 0 | 0 | 0 | 0 |
| Big5_N | 0 | 0 | 0 | 0 | 0 | Big5_N | 0 | 0 | 0 | 0 | 0 |
| Big5_O | 0 | 0 | 0 | 0 | 0 | Big5_O | 0 | 0 | 0 | 0 | 0 |
| SVO_P | 0.015 | 0 | 0 | 0 | 0 | SVO_P | 0 | 0 | 0 | 0 | 0 |
| SVO_I | 0 | 0 | 0 | 0 | 0 | SVO_I | 0 | 0.0094 | 0 | 0 | 0 |
| IRI_F | 0 | 0 | 0 | 0.016 | 0 | IRI_F | 0 | 0 | 0 | 0 | 0 |
| IRI_PT | 0 | 0 | 0 | 0 | 0 | IRI_PT | 0 | 0 | 0 | 0 | 0 |
| IRI_EC | 0 | 0 | 0 | 0 | 0 | IRI_EC | 0 | 0 | 0 | 0 | 0 |
| IRI_PD | 0 | 0 | 0 | 0.0051 | 0 | IRI_PD | 0 | 0 | 0 | 0 | 0 |
| SHS | 0 | 0 | 0.15 | 0 | 0 | SHS | 0 | 0 | 0 | 0 | 0 |
| IQ | 0.079 | 0 | 0 | 0 | -0.015 | IQ | 0 | 0 | 0 | 0 | 0 |
| MVS | 0 | 0 | 0.012 | 0 | 0 | MVS | 0 | 0 | 0 | 0 | 0 |
| PSS | 0 | 0 | 0 | 0 | -0.012 | PSS | 0 | 0 | 0 | 0 | 0 |
| RSS | 0 | 0.034 | 0.020 | 0 | 0 | RSS | 0 | 0 | 0 | 0 | 0 |
| STAI_S | -0.031 | -0.074 | 0 | 0 | 0 | STAI_S | 0 | 0 | 0 | 0 | 0 |
| STAI_T | 0 | 0 | 0 | 0 | -0.039 | STAI_T | 0 | 0 | 0 | 0 | 0 |
| SES | 0.12 | 0.096 | 0 | 0 | 0 | SES | -0.040 | 0 | 0 | 0 | 0 |
| UD | 0 |  |  |  |  | UD | 0 |  |  |  |  |
| INF | 0 |  |  |  |  | INF | 0 |  |  |  |  |
| WP | 0.053 |  |  |  |  | WP | 0 |  |  |  |  |
| area | 0 |  |  |  |  | area | -0.021 |  |  |  |  |
| income | 0.0037 |  |  |  |  | income | -3.1x10<sup>-5</sup> |  |  |  |  |

## Discussion

In this study, we asked a large Japanese sample (N = 6232) to read one of 9 different messages about COVID-19 vaccination and to report their degrees of willingness and refusal to receive the COVID-19 vaccination. We also collected 17 social personality trait scores and demographic information. We analyzed differences in the willingness and refusal to be vaccinated and the underlying psychological processes in an age- and gender-dependent manner.

The overall vaccine acceptance rate was 68.6%, a comparable value with a previous report in Japan [21], although the acceptance rate has been reported to vary in different countries, regions, and the period of time of the data collection [22]. We confirmed that factors including being female, younger age, lower income, no medical problems, and a non-medical workplace were associated with lower motivation to be vaccinated, which is consistent with previous studies [2,16,21]. Importantly, the present study revealed that males in their 10-20s are less motivated to be vaccinated than other groups, and also that prosocial and empathetic consideration, such as agreeableness, fairness, and empathetic concerns, is the driving force of the willingness in younger age groups, while in older people, risk aversion and self-interest are also involved. We also conducted LASSO regression to investigate the social personality traits that motivate COVID-19 vaccination in an age- and gender-dependent manner and found that agreeableness, willingness to protect others and the risk perception of infection generally contribute to the motivation for vaccination in a similar manner to previous studies [15,23]. For males in their 10-20s, who were the least motivated for vaccination, agreeableness followed by inequity aversion and prosocial orientation (preference of joint benefits and dislike of inequity) in social value orientation [24,25,26] contributed to the motivation (Table 3). Furthermore, by evaluating the effectiveness of 9 different messages to promote vaccination, we found that emphasizing the majority’s positive attitudes towards COVID-19 vaccination plus the scientific evidence of safety (i.e., nudge message 8) is most effective for young groups, particularly males in their 10-20s. Although the effect size of this message is not very high, this report provides the first identification of an effective message for young people. This group is an especially important target for vaccinations, because it is likely to be asymptomatic and social, and thus can enlarge the infected population unwittingly.

It was reported that nudges are effective at promoting COVID-19 prevention measures [14,27] and that vaccination motivation was promoted by nudge interventions [28]. As a general strategy of nudges, factors including framing, tone and sender of the message are known to influence whether the reader understands and is motivated by the message [29,30]. However, it was also reported that different nudges made little difference in the context of vaccinations [31]. It is therefore important to clarify why the message emphasizing “majority’s intention to vaccinate + scientific evidence of safety” is effective for young groups, particularly males in their 10-20s, in the present study.

This message apparently appeals to the human bias to conform to social norm [32,33], consistent with other nudge intervention research [34]. However, our analysis of psychological processes demonstrated that the willingness of males in their 10-20s to vaccinate comes from their prosocial and empathetic inclination rather than an obeyance to social rules and norms (consider that neither Big5_C nor ES_S contribute to the willingness to be vaccinated in younger groups). Therefore, it is more likely that the herding effect arising from the “majority’s intention to COVID-19 vaccination + scientific evidence of safety” effectively nudged young people to exhibit their prosocial nature. It is also noteworthy that neither strong discomfort nor resentment was reported from those who received this message (Supplementary Fig. S1 and Table S2 online).

It is puzzling that the effect of nudge message 1 (altruism) was less effective than nudge message 8 (majority + scientific evidence), considering that altruism and agreeableness were the strongest motivators for young people’s inoculation. Head-off effects explain that nudges sometimes do not work on people who have already enough preventive attitudes [35]. Therefore, one possible interpretation of our findings is that the emphasis on the majority’s intention to vaccinate and scientific evidence of safety is more effective than the altruistic message due to head-off effects in the context of COVID-19 vaccination. Related to this, indirectness of the altruistic message may also explain this result. That is, the onset protection effect of the vaccine was established scientifically, but the prevention effect of infection was still being discussed. Therefore, we could only imply in nudge 1 that the self-prevention of infection by vaccination helps others by sparing the number of hospital beds, rather than directly protecting the other’s onset.

For males in their 30s, both agreeableness and stress contributed to the willingness, but for males in their 40s, empathy promoted vaccination, while risk aversion contributed to refusal, indicating that their decision-making balances social benefits and their own risk of the vaccination. For males in their 50s, in addition to empathy and agreeableness, trust and subjective socioeconomic status played a major role, suggesting that this group is sensitive to the expectation of society when making decisions, i.e., social norm, as higher subjective socioeconomic status promoted stronger motivation to vaccinate. The relatively high willingness of this last group to vaccinate in response to nudge message 8 (Fig. 2a) is likely to reflect at least partly their inclination to conform to others (i.e., social norm pressure), as proposed by Sasaki and colleagues [36], making a contrast with people in their 10-20s.

Higher IQ was associated with a greater willingness to vaccinate in females. Related to this, it was reported that higher education is linked to a greater willingness to vaccinate [22]. Our results suggest that this tendency may be stronger in females. For females in their 10-20s, 30s, and 40s, the degree of perceiving others as fair (TRU_WVS), agreeableness, and altruism, respectively, contributed to the willingness to vaccinate, indicating the general tendency that the contribution of prosocial inclination is stronger in females. For females in their 50s, the contribution of empathy became stronger. Females overall, especially in their 40s were less willing to vaccinate than males (Fig. 1a), but at the same time, they were not actively refusing vaccination (Fig. 1b). We think that their lower willingness to vaccination is attributable to differences in lifestyle and social role between genders.

We also found that nudge 2 was the least effective at promoting vaccination motivation. This result is consistent with a previous report that found emphasizing one’s own benefits is less effective than emphasizing public benefits [14]. Interestingly, our post-experimental questionnaire also revealed that nudge 2 was the least likely to induce a sense of responsibility (Supplementary Fig. S1 online).

There are limitations in the present study. First, we could not examine the actual vaccination behaviors of people who were nudged. It was reported that an increase in motivation does not always lead to an actual change in behaviors [37,38,39]. Whether motivation causes behavioral change is an important topic for future investigation. Second, although we identified an effective nudge message for young people with prosocial and empathetic orientation, we still do not have an effective way to nudge young people who have proself orientation. This too is worth future study. The third limitation is cultural differences regarding the majority. Because higher in-group conformity is known for Japanese people [40], it is necessary to examine whether messages emphasizing the majority is generally effective for young people in other cultures. Fourth, we did not include a direct question about the degree to which the participants perceived themselves to be at risk for COVID-19, although the perception of risk was reported as an important factor for determining vaccination attitudes [1,8,9]. We believe that the usual preventive attitudes against COVID-19 reflected risk perceptions and were strongly related to a willingness or refusal to be vaccinated (Tables 1 and 2). Fifth, we did not directly assess the trust of the information source about vaccines or the vaccines themselves. It was reported that this trust affects the willingness to be vaccinated [16,41]. However, we believe that the contribution of risk aversion and loss aversion to the refusal to vaccinate reflects low trust in the information source, at least partly, which rules out the possibility that failure to include trust largely affected the results of the present paper. Last, but not least, although we examined gender differences, we emphasize our findings do not reflect biological differences, but rather cultural and societal differences related to gender. Despite these limitations, the age- and gender-dependent differences in attitudes towards COVID-19 vaccination and the effects of nudge messages we report in this study should provide useful insights into the promotion of COVID-19 vaccination, particularly for young people who are less motivated to be vaccinated.

## Methods

### Data Collection

The survey of willingness to vaccinate and personality traits was conducted between 23 February 2021 and 1 March 2021 (the end of the 3rd wave in Japan). The participants were recruited from the registrants of an internet research service company (NTT Com Research, Inc.).

Among the recruited participants (n = 9,868), 6,332 participants who answered all the questions were paid NTT Com Research points equivalent to $5. No participant was excluded from the analysis. This survey was conducted among people 15-59 years old (male : female = 3409:2823, age = 39.76±11.97). The NICT ethics committee approved all experimental procedures and informed consent was obtained from all participants.

### Questionnaires for personality traits and risk perception of infection

To examine the participants’ social personality traits and items, we utilized the following questionnaires (see Supplementary Table S1 online): loss aversion (LAR), risk aversion (RA), time discounting (TIM), conditional cooperation (CCP), trust (TRU-GSSWVS, TRU-WVS, TRU-GSS; GSS and WVS represent the American General Social Survey, and the World Values Survey, respectively. These three questionnaires were slightly different. That is, GSSWVS, WVS and GSS focused on trustworthiness, fairness and altruism, respectively.), Guilt and Inequity aversions (guilt, inequity, see [42]), Empathizing-Systemizing theory (ES_E,ES_S), Social value orientation (SVO_P, SVO_I,SVO_C), empathy (IRI_F, IRI_PT, IRI_EC, IRI_PD), happiness (SHS), Machiavellianism (MVS), stress (PSS), self-esteem (RSS), anxiety (STAI_S, STAI_T), subjective socioeconomic status (SES), IQ (short version of Leven IQ test), and multi-dimensional characters (Big Five Inventory; Big5_E, Big5_A, Big5_C, Big5_N,Big5_O). As the infection risk index, we asked the participants whether or not they had any underlying medical disease (UD), infected close relatives (INF) or worked in a medical-related place (WP).

We also collected sociodemographic characteristics including age (10-20s, 30s, 40s, and 50s), gender (male:1, female:-1), place of residence (dichotomous variables whether or not the number of the new infections in the living prefecture exceeded the national average during the data acquisition period) and annual income (extremely low; less than $13,000, low; $13,000-$26,000, middle; $26,000-$69,000, high; more than $69,000).

### Assessment of willingness or refusal to be vaccinated

To assess the willingness and refusal to vaccinate, we asked the participants how likely they will get vaccinated for COVID-19 when a vaccine is available (Question A (willingness): do you think you will be vaccinated against the new coronaviruses in the future?) and when to get vaccinated (Question B (refusal): when do you think you will be vaccinated against the new coronaviruses?) Seven response options were provided for each question: 1) not at all, 2) hardly, 3) not much, 4) neither, 5) somewhat, 6) much, and 7) definitely for Question A and 1) as soon as possible, 2) six months later, 3) one year later, 4) two years later, 5) three years later, 6) five years later, and 7) never for Question B.

Participants’ attitudes were examined by the frequencies and ratios summarized in Table 1. We used the chi-square test to evaluate the univariate associations of participants’ attitudes. Regarding the participants’ willingness to vaccinate (Question A), if a participant responded 1) not at all to 3) not much, we judged that the participant had a low likelihood of getting the vaccine. On the other hand, if a participant responded 5) somewhat to 7) definitely, we judged that the participant had a high likelihood of getting the vaccine. Regarding the participants’ refusal to vaccinate (Question B), if a participant responded 1) as soon as possible to 3) one year ahead, we judged that the participant wanted the vaccination immediately. If a participant responded 4) two years to 6) five years later, we judged that the participant intended to be vaccinated eventually. Finally, if a participant responded 7) never, we judged that the participant will always refuse the vaccination. We conducted a general linear model regression analysis of the responses to Question A and Question B to estimate the association with sociodemographic characteristics (Table 2).

### Data analysis

#### Association with personality traits

In order to identify the personality traits that contribute to vaccination intention depending on age and gender, LASSO regression was conducted. As the explanatory variables, we used all the social personality trait scores, the risk index described above, income and place of residence (Table 3) and a dummy binary variable representing the corresponding age and gender. We also included the interactions of the social personality scores with the dummy binary variable. Preventive attitude was not included in the explanatory variables, because we concluded they were affected by the personality traits. LASSO regression [43] was adopted, because the inclusion of interactions increased the dimensionality of explanatory variables, which makes the interpretation of the regression results easier, as the effective explanatory variables were sparsely selected and non-effective explanatory variables were set to 0. The regression function was in the form of

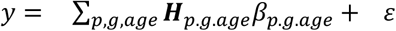

where *y* = [*y*_1_,…,*y*_*n*_]^*T*^ is the responses to Question A (willingness to be vaccinated) or the binary transformed responses to Question B (representing refuse (7) or not refuse (1-6) to assess the refusal to the vaccination).

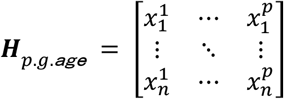

is the matrix with each personality score (p) of each participant (n) of each gender (m, males; f, females) and age. 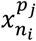 was the product of the score of the personality (j) and a dummy variable that was set to 1 if participant (i) belonged to the gender (g) and age group. The parameters 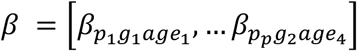 were computed as a solution of

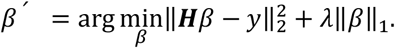

The Euclidean norm of the first term expressed the least square approximation of the regression component, while the second term with ||±||_1_ = Σ |±_*p.g.age*_| was used to produce a sparse representation. A larger *λ* indicates a larger weight on the minimization of ||±||_1_ and thus on sparseness. The optimal value of *λ* was determined by 10-fold cross validation using cv.glmnet [44] function of R (a language and environment for statistical computing and graphics similar to S, http://cran.r-project.org/.).

#### Evaluating age- and gender-dependent effects of nudges

Messages emphasizing social benefits have been reported to increase the willingness to receive the influenza vaccine [45] as well as the willingness to cooperate in the prevention of the spread of COVID-19 [15,36]. With reference to previous reports, a message emphasizing altruism was used as the control nudge (nudge 1, see Appendix 1). In addition, based on several reports indicating that scientific evidence, such as information about vaccine efficacy and side effects, influences the motivation for COVID-19 vaccination [12,46], we included scientific information in the messages.

The framing effect concerning a loss or gain representation of the same issue is known to affect the nudge [35,47,48,49]. Therefore, we included both loss and gain frames for altruism and scientific evidence. Importantly, it was reported that the majority also affects vaccination motivation, and Sasaki and colleagues suggested that the motivation to vaccinate increases when the vaccination rate of the same age-group is high [36]. Therefore, we also included messages that emphasized the majority of others.

Each participant read one of the 9 nudge messages once before being asked their willingness and refusal to vaccinate. All nudges were given to almost equal numbers of participants in terms of age and gender. The numbers of participants assigned to nudge messages 1 to 9 were 705 (323), 695 (311), 684 (308), 689 (310), 711 (323), 699 (318), 695 (308), 677 (310), and 687 (312), respectively, with the number of females in brackets. To assess the age- and gender-dependent impact on the willingness to vaccinate, we compared the ratios of the participants who were strongly motivated to vaccinate (i.e., Question A, 6 or 7) between the control nudge (nudge 1) and other nudges in each age and gender group. To assess the effect to reduce the refusal to vaccinate, the ratios of people who rejected vaccination (i.e., Question B, 7) between the control nudge (nudge 1) and other nudges were compared. A function of R, prop.test, was used to test the difference in proportions, with the significance level of a one-tailed p = 0.05.

We conducted LASSO regression, which included the interactions of social personality trait scores with age, gender or the dummy variables representing which message was sent to each participant, in addition to social personality trait scores, demographic information, annual income and place of residence, and any UD, INF and WP.

To directly assess the personality traits and age which nudge message 8 influenced, we conducted LASSO regression only for participants who received nudge 8 (n = 677). The 17 personalities and their interactions with binary age group variables were used as regressors. The demographic information, annual income, place of residence, UD, INF and WP were also included as regressors.

#### Post experimental questionnaire

We finally requested the participants to evaluate the nudge message they received after being asked to answer their willingness and refusal to COVID-19 vaccination. We asked them to answer the following 7 statements on a 5-point scale ranging from true to false: “I feel responsible”, “I feel empathy”, “I feel peer pressure”, “I feel repulsed”, “I feel uncomfortable”, “I find it very stimulating” and “It seems memorable”.

## Data Availability

The datasets analyzed during the current study are available from the corresponding author upon reasonable request.

## Acknowledgements

The authors disclose receipt of the following financial support for the research, authorship, and/or publication of this article: Preparation of this manuscript was supported by CREST, JST, Moonshot R&D Grant Number JPMJMS2011, JST and KAKENNH (17H06314 and 20K21562) to M.H. We would like to thank Satoshi Tada and Koji Fuji for technical assistance.

## Author Contribution

MH, TN and FO designed the study. TT and MH performed the research and analyzed the data. TT wrote the first draft and MH edited it. TT, TN, FO, and MH wrote the paper.

## Additional information

The authors declare no competing interests.

## Appendix1

Short messages (nudges) for promoting vaccination used in the experiments. (English translations are followed by the original Japanese.)

Description.

Vaccination is considered effective for preventing the spread of COVID-19.

1. control (altruism; gain framing) Your vaccination will help spare the number of hospital beds and save people’s lives.
2. scientific evidence (self-interest; gain framing) A report in the U.S. scientific journal Science showed that one in 60,000 people developed acute severe allergic symptoms after vaccination and that vaccination reduced the number of people with COVID-19 by one-twentieth.
3. scientific evidence (self-interest; loss framing) A report in the U.S. scientific journal Science showed that one in 60,000 people develop acute severe allergic symptoms after vaccination and that the number of people who develop COVID-19 increases 20-fold without vaccination.
4. scientific evidence (self-interest; gain framing) + altruism (gain framing) A report in the U.S. scientific journal Science showed that 1 in 60,000 people developed acute severe allergic symptoms after vaccination and that vaccination reduced the number of people with COVID-19 by a factor of 20. Your vaccination will lead to more room in hospital beds and save lives.
5. scientific evidence (self-interest; loss framing) + altruism (gain framing) A report in the U.S. scientific journal Science showed that 1 in 60,000 people showed acute severe allergic symptoms after vaccination and that the number of people who develop COVID-19 increases 20-fold without vaccination. Your vaccination will help spare hospital beds and save lives.
6. scientific evidence (self-interest; gain framing) + altruism (loss framing) A report in the U.S. scientific journal Science showed that one in 60,000 people showed acute severe allergic symptoms after vaccination and that vaccination reduces the number of people with COVID-19 by one-twentieth. If you do not vaccinate, there will be a shortage of hospital beds and people’s lives will be at risk.
7. scientific evidence (self-interest; loss framing) + altruism (loss framing) A report in the U.S. scientific journal Science showed that 1 in 60,000 people showed acute severe allergic symptoms after vaccination and that the number of people who develop COVID-19 increases 20-fold if they are not vaccinated. If you do not vaccinate, there will be a shortage of hospital beds and people’s lives will be at risk.
8. scientific evidence (self-interest; gain framing) + majority A report in the U.S. scientific journal Science showed that only one in 60,000 people showed acute severe allergic symptoms after vaccination and that vaccination reduced the number of people with COVID-19 by one-twentieth. According to a survey by Ipsos, a global research firm, about 70% of people agree to be vaccinated.
9. scientific evidence (self-interest; loss framing) + majority A report in the U.S. scientific journal Science showed that one in 60,000 people showed acute severe allergic symptoms after vaccination and that the number of people who develop COVID-19 increases 20-fold without vaccination. According to a survey by Ipsos, a global research firm, about 70 percent of people agree to be vaccinated.

