## Supplemental files for "Age- and gender-dependent differences in attitudes towards COVID-19 vaccination and underlying psychological processes"

#### Abstract

The most promising way to prevent the explosive spread of COVID-19 infection is to achieve herd immunity through vaccination. It is therefore important to motivate those who are less willing to be vaccinated. To address this issue, we conducted an online survey of 6232 Japanese people to investigate age- and gender- dependent differences in attitudes towards COVID-19 vaccination and the underlying psychological processes. We asked participants to read one of nine different messages about COVID-19 vaccination and rate their willingness to be vaccinated. We also collected their 17 social personality trait scores and demographic information. We found that males 10-20 years old showed the minimum willingness to be vaccinated. We also found that prosocial traits are the driving force for young people, but the motivation in older people also depends on risk aversion and self-interest. Furthermore, an analysis of 9 different messages demonstrated that for young people (particularly males), the message emphasizing the majority's intention to vaccinate and scientific evidence for the safety of the vaccination had the strongest positive effect on the willingness to be vaccinated, suggesting that the herding effect arising from the "majority + scientific evidence" message nudges young people to show their prosocial nature in action.

Figure S1. Subjective evaluation of messages.

The percentage of people who scored in each of the seven subjective ratings (responsibleness, empathy, peer pressure, repulsed, uncomfortable, stimulating, and memorable) by gender and age are shown. Feelings of responsibility were lower among men in their 10-20s in response to nudge 2 (p = 4.9 x$10^{-3}$). No significantly greater senses of repulsion or discomfort occurred in response to nudge 8. Significant differences compared with control (nudge 1) are indicated by the asterisks (*). The p-values of the black asterisks for feeling responsible are, from left (male) to right (female), 4.9 x $10^{-3}$, 0.043, 7.6 x $10^{-3}$, and 0.031, and those of the blue asterisks are 0.012, 0.024, 0.011, and 0.021; for empathy, they are 0.038, 0.035, 0.027, and 0.013 and 0.041 and 0.022, respectively; for peer pressure, they are 0.021 and 0.019 and 9.4 x$10^{-3}$, 0.010, and 0.044, respectively; for repulsion, they are 0.031 and 0.036, respectively; for discomfort, they are 0.047 and 0.030, 0.049, and 0.028, respectively; for stimulating, they are 0.036, 0.033, and 7.1 x $10^{-3}$ and 7.4 x $10^{-3}$, 0.011, 0.037, and 0.022, respectively; and for feeling memorable, they are 0.019, 0.045, and 0.032.

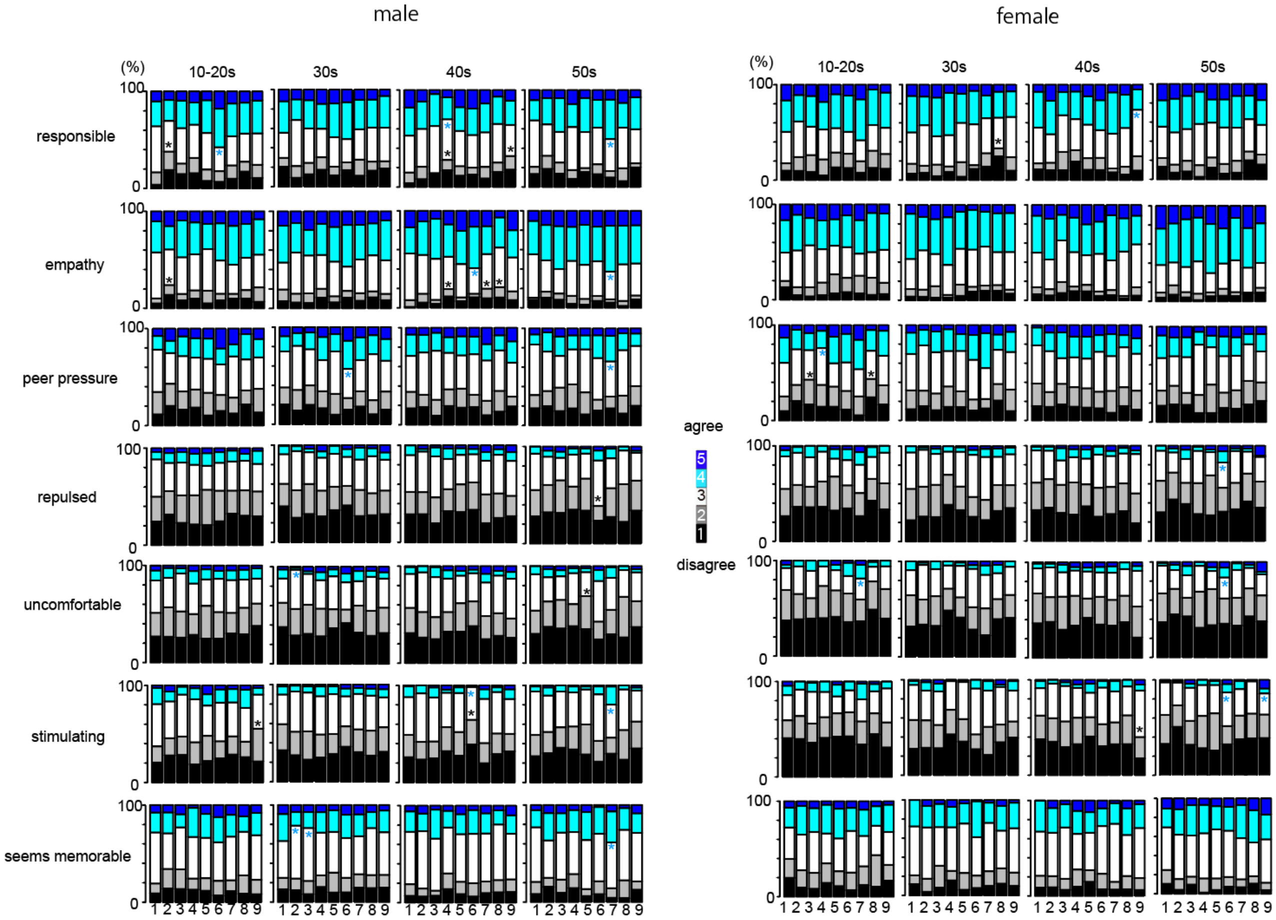

Table S1. Detailed list of the personality tests used in this study.

| Abbreviation | subscore abbreviation (item number) | Questionnaire name | references |
|---|---|---|---|
| Big5 | | Big Five personality traits | 1,2 |
| Extraversion | Big5_E (12) | | |
| Agreeableness | Big5_A (12) | | |
| Consciousness | Big5_C (12) | | |
| Neuroticism | Big5_N (12) | | |
| Openness | Big5_O (12) | | |
| IRI | | Interpersonal Reactivity Index | 3,4 |
| Fantasy | IRI_F (7) | | |
| Perspective taking | IRI_PT (7) | | |
| Empathic Concern | IRI_EC (7) | | |
| Personal Distress | IRI_PD (7) | | |
| MVS | (20) | Machiavellianism Scale | 5,6 |
| PSS | (10) | Perceived Subjective Stress | 7,8 |
| RA | (10) | Risk Aversion | 9,10 |
| RSS | (10) | Rosenberg Self - Esteem scale | 11,12 |
| SES | (1) | Socio Economic Status | 13,14 |
| SVO | | Social Value Orientation | 15,16 |
| Prosocial | SVO_P (8) | | |
| indvidualist | SVO_I (8) | | |
| Competitor | SVO_C (8) | | |
| STAI | | State - Trait Anxiety Inventory | 17,18 |
| State | STAI_S (20) | | |
| trait | STAI_T (20) | | |
| TIM | (15) | Time discounting | 19,20 |
| CCP | (1) | conditional cooperation | 21 |
| TRU | | | |
| trust | TRU_GSSWVS (1) | the World Values survey/the American General Social Survey | |
| fairness | TRU_WVS (1) | the World Values survey | |
| altruism | TRU_GSS (1) | the American General Social Survey | |
| LAR | loss aversion (6) | lottery choice task | 22 |
| Inequity Guilt aversion | | | 27 |
| guilt-aversion | guilt | | |
| inequity-aversion | inequity | | |
| ES | | Empathizing–Systemizing theory | 23 |
| Systemizing | ES_S (20) | | |
| Empathizing | ES_E (20) | | |
| SHS | (4) | Tendency to give socially desirable answers | 24,25 |
| IQ | (9) | Raven Advanced Progressive Matrices test #11,24,28,36,43,48,49,53,55 | 26 |

### Table S2. Subjective evaluation of the messages.

| | | all | | | | | | | | | male | | | | | | | | | female | | | | | | | | |
|---|---|---|---|---|---|---|---|---|---|---|---|---|---|---|---|---|---|---|---|---|---|---|---|---|---|---|---|---|
| items | nudge | total | disagree (12) | | nutral (3) | | agree (45) | | F/chi_sq | p_value | total | disagree (12) | | nutral (3) | | agree (45) | | F/chi_sq | p_value | total | disagree (12) | | nutral (3) | | agree (45) | | F/chi_sq | p_value |
| | | n | n | % | n | % | n | % | | | n | n | % | n | % | n | % | | | n | n | % | n | % | n | % | | |
| | | 6232 | | | | | | | | | 3409 | | | | | | | | | 2823 | | | | | | | | |
| | total | | 1285 | 20.6 | 2180 | 35.0 | 2627 | 42.2 | | | | 806 | 23.6 | 1216 | 35.7 | 1387 | 40.7 | | | | 619 | 21.9 | 964 | 34.1 | 1240 | 43.9 | | |
| | 1 | 705 | 140 | 19.9 | 259 | 36.7 | 306 | 43.4 | 29.517 | 0.0207 | 382 | 80 | 20.9 | 149 | 39.0 | 153 | 40.1 | 30.59 | 0.0152 | 323 | 60 | 18.6 | 110 | 34.1 | 153 | 47.4 | 19.29 | 0.254 |
| | 2 | 695 | 162 | 23.3 | 258 | 37.1 | 275 | 39.6 | | | 384 | 105 | 27.3 | 146 | 38.0 | 133 | 34.6 | | | 311 | 57 | 18.3 | 112 | 36.0 | 142 | 45.7 | | |
| | 3 | 684 | 155 | 22.7 | 244 | 35.7 | 285 | 41.7 | | | 376 | 84 | 22.3 | 143 | 38.0 | 149 | 39.6 | | | 308 | 71 | 23.1 | 101 | 32.8 | 136 | 44.2 | | |
| | 4 | 689 | 163 | 23.7 | 246 | 35.7 | 270 | 39.2 | | | 369 | 98 | 26.6 | 138 | 37.4 | 133 | 36.0 | | | 310 | 65 | 21.0 | 108 | 34.8 | 137 | 44.2 | | |
| responsible | 5 | 711 | 155 | 21.8 | 247 | 34.7 | 309 | 43.5 | | | 388 | 79 | 20.4 | 136 | 35.1 | 173 | 44.6 | | | 323 | 76 | 23.5 | 111 | 34.4 | 136 | 42.1 | | |
| | 6 | 699 | 150 | 21.5 | 220 | 31.5 | 329 | 47.1 | | | 381 | 83 | 21.8 | 116 | 30.4 | 182 | 47.8 | | | 318 | 67 | 21.1 | 104 | 32.7 | 147 | 46.2 | | |
| | 7 | 695 | 148 | 21.3 | 224 | 32.2 | 323 | 46.5 | | | 387 | 87 | 22.5 | 126 | 32.6 | 174 | 45.0 | | | 308 | 61 | 19.8 | 98 | 31.8 | 149 | 48.4 | | |
| | 8 | 677 | 171 | 25.3 | 240 | 35.5 | 266 | 39.3 | | | 367 | 89 | 24.3 | 136 | 37.1 | 142 | 38.7 | | | 310 | 82 | 26.5 | 104 | 33.5 | 124 | 40.0 | | |
| | 9 | 687 | 181 | 26.3 | 242 | 35.2 | 264 | 38.4 | | | 375 | 101 | 26.9 | 126 | 33.6 | 148 | 39.5 | | | 312 | 80 | 25.6 | 116 | 37.2 | 116 | 37.2 | | |
| | total | | 918 | 14.7 | 2170 | 34.8 | 3144 | 50.4 | | | | 511 | 15.0 | 1218 | 35.7 | 1680 | 49.3 | | | | 407 | 14.4 | 952 | 33.7 | 1464 | 51.9 | | |
| | 1 | 705 | 92 | 13.0 | 260 | 36.9 | 353 | 50.1 | 10.85 | 0.82 | 382 | 47 | 12.3 | 158 | 41.4 | 177 | 46.3 | 24.39 | 0.0814 | 323 | 45 | 13.9 | 102 | 31.6 | 176 | 54.5 | 11.06 | 0.806 |
| | 2 | 695 | 107 | 15.4 | 252 | 36.3 | 336 | 48.3 | | | 384 | 66 | 17.2 | 146 | 38.0 | 172 | 44.8 | | | 311 | 41 | 13.2 | 106 | 34.1 | 164 | 52.7 | | |
| | 3 | 684 | 94 | 13.7 | 251 | 36.7 | 339 | 49.6 | | | 376 | 50 | 13.3 | 136 | 36.2 | 190 | 50.5 | | | 308 | 44 | 14.3 | 115 | 37.3 | 149 | 48.4 | | |
| | 4 | 689 | 102 | 14.8 | 236 | 34.3 | 341 | 49.5 | | | 369 | 65 | 17.6 | 130 | 35.2 | 174 | 47.2 | | | 310 | 37 | 11.9 | 106 | 34.2 | 167 | 53.9 | | |
| empathy | 5 | 711 | 105 | 14.8 | 239 | 33.6 | 367 | 51.6 | | | 388 | 48 | 12.4 | 142 | 36.6 | 198 | 51.0 | | | 323 | 57 | 17.6 | 97 | 30.0 | 169 | 52.3 | | |
| | 6 | 699 | 105 | 15.0 | 224 | 32.0 | 370 | 52.9 | | | 381 | 57 | 15.0 | 114 | 29.9 | 210 | 55.1 | | | 318 | 48 | 15.1 | 110 | 34.6 | 160 | 50.3 | | |
| | 7 | 695 | 107 | 15.4 | 223 | 32.1 | 365 | 52.5 | | | 387 | 58 | 15.0 | 126 | 32.6 | 203 | 52.5 | | | 308 | 49 | 15.9 | 97 | 31.5 | 162 | 52.6 | | |
| | 8 | 677 | 100 | 14.8 | 244 | 36.0 | 333 | 49.2 | | | 367 | 59 | 16.1 | 138 | 37.6 | 170 | 46.3 | | | 310 | 41 | 13.2 | 106 | 34.2 | 163 | 52.6 | | |
| | 9 | 687 | 106 | 15.4 | 241 | 35.1 | 340 | 49.5 | | | 375 | 61 | 16.3 | 128 | 34.1 | 186 | 49.6 | | | 312 | 45 | 14.4 | 113 | 36.2 | 154 | 49.4 | | |
| | total | | 2088 | 33.5 | 2351 | 37.7 | 1793 | 28.8 | | | | 1141 | 33.5 | 1324 | 38.8 | 944 | 27.7 | | | | 947 | 33.5 | 1027 | 36.4 | 849 | 30.1 | | |
| | 1 | 705 | 235 | 33.3 | 280 | 39.7 | 190 | 27.0 | 45.00 | 1.39x10-4 | 382 | 132 | 34.6 | 161 | 42.1 | 89 | 23.3 | 30.68 | 0.015 | 323 | 103 | 31.9 | 119 | 36.8 | 101 | 31.3 | 30.88 | 0.014 |
| | 2 | 695 | 243 | 35.0 | 281 | 40.4 | 171 | 24.6 | | | 384 | 137 | 35.7 | 157 | 40.9 | 90 | 23.4 | | | 311 | 106 | 34.1 | 124 | 39.9 | 81 | 26.0 | | |
| | 3 | 684 | 241 | 35.2 | 254 | 37.1 | 189 | 27.6 | | | 376 | 132 | 35.1 | 148 | 39.4 | 96 | 25.5 | | | 308 | 109 | 35.4 | 106 | 34.4 | 93 | 30.2 | | |
| | 4 | 689 | 244 | 35.4 | 251 | 36.4 | 184 | 26.7 | | | 369 | 138 | 37.4 | 128 | 34.7 | 103 | 27.9 | | | 310 | 106 | 34.2 | 123 | 39.7 | 81 | 26.1 | | |
| peer pressure | 5 | 711 | 244 | 34.3 | 264 | 37.1 | 203 | 28.6 | | | 388 | 132 | 34.0 | 154 | 39.7 | 102 | 26.3 | | | 323 | 112 | 34.7 | 110 | 34.1 | 101 | 31.3 | | |
| | 6 | 699 | 201 | 28.8 | 261 | 37.3 | 237 | 33.9 | | | 381 | 110 | 28.9 | 141 | 37.0 | 130 | 34.1 | | | 318 | 91 | 28.6 | 120 | 37.7 | 107 | 33.6 | | |
| | 7 | 695 | 200 | 28.8 | 248 | 35.7 | 247 | 35.5 | | | 387 | 114 | 29.5 | 146 | 37.7 | 127 | 32.8 | | | 308 | 86 | 27.9 | 102 | 33.1 | 120 | 39.0 | | |
| | 8 | 677 | 238 | 35.2 | 270 | 39.9 | 169 | 25.0 | | | 367 | 115 | 31.3 | 157 | 42.8 | 95 | 25.9 | | | 310 | 123 | 39.7 | 113 | 36.5 | 74 | 23.9 | | |
| | 9 | 687 | 242 | 35.2 | 242 | 35.2 | 203 | 29.5 | | | 375 | 131 | 34.9 | 132 | 35.2 | 112 | 29.9 | | | 312 | 111 | 35.6 | 110 | 35.3 | 91 | 29.2 | | |
| | total | | 3569 | 6.3864 | 2005 | 289.1 | 655 | 94.458 | | | | 1889 | 55.4 | 1134 | 33.3 | 386 | 11.3 | | | | 1683 | 59.6 | 871 | 30.9 | 269 | 9.5 | | |
| | 1 | 705 | 398 | 56.5 | 248.0 | 35.2 | 59 | 8.4 | 32.52 | 8.55x10-3 | 382 | 209 | 54.7 | 137 | 35.9 | 36 | 9.4 | 17 | 0.386 | 323 | 189 | 58.5 | 111 | 34.4 | 23 | 7.1 | 31.25 | 0.0125 |
| | 2 | 695 | 408 | 58.7 | 234.0 | 33.7 | 53 | 7.6 | | | 384 | 212 | 55.2 | 137 | 35.7 | 35 | 9.1 | | | 311 | 196 | 63.0 | 97 | 31.2 | 18 | 5.8 | | |
| | 3 | 684 | 393 | 57.5 | 212.0 | 31.0 | 79 | 11.5 | | | 376 | 211 | 56.1 | 124 | 33.0 | 41 | 10.9 | | | 308 | 182 | 59.1 | 88 | 28.6 | 38 | 12.3 | | |
| | 4 | 689 | 399 | 57.9 | 210.0 | 30.5 | 70 | 10.2 | | | 369 | 202 | 54.7 | 120 | 32.5 | 47 | 12.7 | | | 310 | 197 | 63.5 | 90 | 29.0 | 23 | 7.4 | | |
| repulsed | 5 | 711 | 436 | 61.3 | 199.0 | 28.0 | 73 | 10.3 | | | 388 | 234 | 60.3 | 113 | 29.1 | 41 | 10.6 | | | 323 | 205 | 63.5 | 86 | 26.6 | 32 | 9.9 | | |
| | 6 | 699 | 378 | 54.1 | 234.0 | 33.5 | 87 | 12.4 | | | 381 | 205 | 53.8 | 129 | 33.9 | 47 | 12.3 | | | 318 | 173 | 54.4 | 105 | 33.0 | 40 | 12.6 | | |
| | 7 | 695 | 372 | 53.5 | 230.0 | 33.1 | 93 | 13.4 | | | 387 | 209 | 54.0 | 123 | 31.8 | 55 | 14.2 | | | 308 | 163 | 52.9 | 107 | 34.7 | 38 | 12.3 | | |
| | 8 | 677 | 397 | 58.6 | 217.0 | 32.1 | 63 | 9.3 | | | 367 | 200 | 54.5 | 133 | 36.2 | 34 | 9.3 | | | 310 | 197 | 63.5 | 84 | 27.1 | 29 | 9.4 | | |
| | 9 | 687 | 388 | 56.5 | 221.0 | 32.2 | 78 | 11.4 | | | 375 | 207 | 55.2 | 118 | 31.5 | 50 | 13.3 | | | 312 | 181 | 58.0 | 103 | 33.0 | 28 | 9.0 | | |
| | total | | 3761 | 60.3 | 1847 | 29.6 | 624 | 10.0 | | | | 1934 | 56.7 | 1101 | 32.3 | 374 | 11.0 | | | | 1827 | 64.7 | 746 | 26.4 | 250 | 8.9 | | |
| | 1 | 705 | 434 | 61.6 | 208.0 | 29.5 | 63 | 8.9 | 35.21 | 3.7x10-3 | 382 | 217 | 56.8 | 124 | 32.5 | 41 | 10.7 | 17.9 | 0.33 | 323 | 217 | 67.2 | 84 | 26.0 | 22 | 6.8 | 30.26 | 0.0167 |
| | 2 | 695 | 422 | 60.7 | 224.0 | 32.2 | 49 | 7.1 | | | 384 | 220 | 57.3 | 133 | 34.6 | 31 | 8.1 | | | 311 | 202 | 65.0 | 91 | 29.3 | 18 | 5.8 | | |
| | 3 | 684 | 407 | 59.5 | 209.0 | 30.6 | 68 | 9.9 | | | 376 | 214 | 56.9 | 127 | 33.8 | 35 | 9.3 | | | 308 | 193 | 62.7 | 82 | 26.6 | 33 | 10.7 | | |
| | 4 | 689 | 421 | 61.1 | 193.0 | 28.0 | 65 | 9.4 | | | 369 | 205 | 55.6 | 117 | 31.7 | 47 | 12.7 | | | 310 | 216 | 69.7 | 76 | 24.5 | 18 | 5.8 | | |
| unconfortable | 5 | 711 | 452 | 63.6 | 194.0 | 27.3 | 65 | 9.1 | | | 388 | 238 | 61.3 | 112 | 28.9 | 38 | 9.8 | | | 323 | 214 | 66.3 | 82 | 25.4 | 27 | 8.4 | | |
| | 6 | 699 | 400 | 57.2 | 210.0 | 30.0 | 89 | 12.7 | | | 381 | 206 | 54.1 | 126 | 33.1 | 49 | 12.9 | | | 318 | 194 | 61.0 | 84 | 26.4 | 40 | 12.6 | | |
| | 7 | 695 | 389 | 56.0 | 208.0 | 29.9 | 98 | 14.1 | | | 387 | 209 | 54.0 | 121 | 31.3 | 57 | 14.7 | | | 308 | 180 | 58.4 | 87 | 28.2 | 41 | 13.3 | | |
| | 8 | 677 | 425 | 62.8 | 193.0 | 28.5 | 59 | 8.7 | | | 367 | 212 | 57.8 | 121 | 33.0 | 34 | 9.3 | | | 310 | 213 | 68.7 | 72 | 23.2 | 25 | 8.1 | | |
| | 9 | 687 | 411 | 59.8 | 208.0 | 30.3 | 68 | 9.9 | | | 375 | 213 | 56.8 | 120 | 32.0 | 42 | 11.2 | | | 312 | 198 | 63.5 | 88 | 28.2 | 26 | 8.3 | | |
| | | | 3403 | 54.605 | 2211 | 35.478 | 618 | 9.9 | | | | 1732 | 50.8 | 1313 | 38.5 | 364 | 10.7 | | | | 1671 | 59.2 | 898 | 31.8 | 254 | 9.0 | | |
| | 1 | 705 | 385 | 54.6 | 250.0 | 35.5 | 70 | 9.9 | 24.626 | 0.077 | 382 | 189 | 49.5 | 149 | 39.0 | 44 | 11.5 | 17.46 | 0.357 | 323 | 196 | 60.7 | 101 | 31.3 | 26 | 8.0 | 26.13 | 0.0523 |
| | 2 | 695 | 380 | 54.7 | 255.0 | 36.7 | 60 | 8.6 | | | 384 | 183 | 47.7 | 162 | 42.2 | 39 | 10.2 | | | 311 | 197 | 63.3 | 93 | 29.9 | 21 | 6.8 | | |
| | 3 | 684 | 358 | 52.3 | 263.0 | 38.5 | 63 | 9.2 | | | 376 | 187 | 49.7 | 155 | 41.2 | 34 | 9.0 | | | 308 | 171 | 55.5 | 108 | 35.1 | 29 | 9.4 | | |
| | 4 | 689 | 387 | 56.2 | 234.0 | 34.0 | 58 | 8.4 | | | 369 | 191 | 51.8 | 141 | 38.2 | 37 | 10.0 | | | 310 | 196 | 63.2 | 93 | 30.0 | 21 | 6.8 | | |
| stimulating | 5 | 711 | 406 | 57.1 | 236.0 | 33.2 | 69 | 9.7 | | | 388 | 203 | 52.3 | 146 | 37.6 | 39 | 10.1 | | | 323 | 203 | 62.8 | 90 | 27.9 | 30 | 9.3 | | |
| | 6 | 699 | 374 | 53.5 | 258.0 | 36.9 | 67 | 9.6 | | | 381 | 201 | 52.8 | 144 | 37.8 | 36 | 9.4 | | | 318 | 173 | 54.4 | 114 | 35.8 | 31 | 9.7 | | |
| | 7 | 695 | 358 | 51.5 | 239.0 | 34.4 | 98 | 14.1 | | | 387 | 183 | 47.3 | 147 | 38.0 | 57 | 14.7 | | | 308 | 175 | 56.8 | 92 | 29.9 | 41 | 13.3 | | |
| | 8 | 677 | 376 | 55.5 | 242.0 | 35.7 | 59 | 8.7 | | | 367 | 185 | 50.4 | 144 | 39.2 | 38 | 10.4 | | | 310 | 191 | 61.6 | 98 | 31.6 | 21 | 6.8 | | |
| | 9 | 687 | 379 | 55.2 | 234.0 | 34.1 | 74 | 10.8 | | | 375 | 210 | 56.0 | 125 | 33.3 | 40 | 10.7 | | | 312 | 169 | 54.2 | 109 | 34.9 | 34 | 10.9 | | |
| | total | | 1475 | 23.7 | 2846 | 45.7 | 1911 | 30.7 | | | | 799 | 23.4 | 1618 | 47.5 | 992 | 29.1 | | | | 676 | 23.9 | 1228 | 43.5 | 919 | 32.6 | | |
| | 1 | 705 | 169 | 24.0 | 330.0 | 46.8 | 206 | 29.2 | 14.89 | 0.53 | 382 | 81 | 21.2 | 190 | 49.7 | 111 | 29.1 | 17.83 | 0.334 | 323 | 88 | 27.2 | 140 | 43.3 | 95 | 29.4 | 13.87 | 0.61 |
| | 2 | 695 | 155 | 22.3 | 327.0 | 47.1 | 213 | 30.6 | | | 384 | 91 | 23.7 | 185 | 48.2 | 108 | 28.1 | | | 311 | 64 | 20.6 | 142 | 45.7 | 105 | 33.8 | | |
| | 3 | 684 | 154 | 22.5 | 329.0 | 48.1 | 201 | 29.4 | | | 376 | 83 | 22.1 | 190 | 50.5 | 103 | 27.4 | | | 308 | 71 | 23.1 | 139 | 45.1 | 98 | 31.8 | | |
| | 4 | 689 | 185 | 26.9 | 288.0 | 41.8 | 206 | 29.9 | | | 369 | 103 | 27.9 | 164 | 44.4 | 102 | 27.6 | | | 310 | 82 | 26.5 | 124 | 40.0 | 104 | 33.5 | | |
| memorable | 5 | 711 | 164 | 23.1 | 320.0 | 45.0 | 227 | 31.9 | | | 388 | 88 | 22.7 | 178 | 45.9 | 122 | 31.4 | | | 323 | 76 | 23.5 | 142 | 44.0 | 105 | 32.5 | | |
| | 6 | 699 | 154 | 22.0 | 319.0 | 45.6 | 226 | 32.3 | | | 381 | 84 | 22.0 | 174 | 45.7 | 123 | 32.3 | | | 318 | 70 | 22.0 | 145 | 45.6 | 103 | 32.4 | | |
| | 7 | 695 | 155 | 22.3 | 315.0 | 45.3 | 225 | 32.4 | | | 387 | 86 | 22.2 | 174 | 45.0 | 127 | 32.8 | | | 308 | 69 | 22.4 | 141 | 45.8 | 98 | 31.8 | | |
| | 8 | 677 | 179 | 26.4 | 304.0 | 44.9 | 194 | 28.7 | | | 367 | 92 | 25.1 | 188 | 51.2 | 87 | 23.7 | | | 310 | 87 | 28.1 | 116 | 37.4 | 107 | 34.5 | | |
| | 9 | 687 | 160 | 23.3 | 314.0 | 45.7 | 213 | 31.0 | | | 375 | 91 | 24.3 | 175 | 46.7 | 109 | 29.1 | | | 312 | 69 | 22.1 | 139 | 44.6 | 104 | 33.3 | | |

17. Spielberger, C. D. *State-Trait Anxiety Inventory: Bibliography.* (Consulting

Psychologists Press, 1989).

26. Jensen, A. R. *The g factor: The science of mental ability.* (Praeger, 1998).

27. Nihonsugi, T., Ihara, A. & Haruno, M. Selective increase of intention-based economic decisions by noninvasive brain stimulation to the dorsolateral prefrontal cortex. *J. Neurosci*. **35**, 3412-3419 (2015).
